# Cardiovascular risk factors are independently associated with COVID-19 mortality: a prospective cohort study

**DOI:** 10.1101/2020.10.01.20205229

**Authors:** Didier Collard, Nick S. Nurmohamed, Yannick Kaiser, Laurens F. Reeskamp, Tom Dormans, Hazra Moeniralam, Suat Simsek, Renée A. Douma, Annet Eerens, Auke C. Reidinga, Paul Elbers, Martijn Beudel, Liffert Vogt, Erik S.G. Stroes, Bert-Jan H. van den Born

**Affiliations:** Amsterdam UMC, University of Amsterdam, Department of Vascular Medicine, Amsterdam Cardiovascular Sciences, Amsterdam, the Netherlands; Amsterdam UMC, Vrije Universiteit Amsterdam, Department of Cardiology, Amsterdam Cardiovascular Sciences, Amsterdam, the Netherlands; Zuyderland Medisch Centrum, Department of Intensive Care, Sittard-Geleen, the Netherlands; St. Antonius Hospital, Department of Internal Medicine, Nieuwegein, the Netherlands; Northwest Clinics, Department of Internal Medicine, Alkmaar, The Netherlands; Flevohospital, Department of Internal Medicine, Almere, The Netherlands; Treant Healthcare Group, Department of Oncology, Hoogeveen, The Netherlands; Martini Hospital, Intensive Care Unit, Groningen, The Netherlands; Amsterdam UMC, Vrije Universiteit Amsterdam, Department of Intensive Care, Amsterdam Amsterdam, the Netherlands; Amsterdam UMC, University of Amsterdam, Department of Neurology, Amsterdam Neuroscience Institute, Amsterdam, the Netherlands; Amsterdam UMC, University of Amsterdam, Department of Nephrology, Amsterdam Cardiovascular Sciences, Amsterdam, the Netherlands; Amsterdam UMC, University of Amsterdam, Department of Public Health, Amsterdam, the Netherlands

**Author notes:** Address for correspondence Dr. Bert-Jan H. van den Born, Department of Vascular Medicine, Amsterdam University Medical Centers, University of Amsterdam, Meibergdreef 9, 1105 AZ Amsterdam, the Netherlands. These authors contributed equally to this work (Didier Collard, Nick S. Nurmohamed, Yannick Kaiser).

**Keywords:** COVID-19, SARS-CoV-2, CVD, hypertension, diabetes, mortality, ICU-admission

## Abstract

**Objectives:** Recent reports suggest a high prevalence of hypertension and diabetes in COVID-19 patients, but the role of cardiovascular disease (CVD) risk factors in the clinical course of COVID-19 is unknown. We evaluated the time-to-event relationship between hypertension, dyslipidemia, diabetes, and COVID-19 outcomes.

**Design:** We analyzed data from the prospective Dutch COVID-PREDICT cohort, an ongoing prospective study of patients admitted for COVID-19 infection.

**Setting:** Patients from 8 participating hospitals, including two university hospitals from the COVID-PREDICT cohort were included.

**Participants:** Admitted, adult patients with a positive COVID-19 polymerase chain reaction (PCR) or high suspicion based on CT-imaging of the thorax. Patients were followed for major outcomes during hospitalization. CVD risk factors were established via home medication lists and divided in antihypertensives, lipid lowering therapy, and antidiabetics.

**Primary and secondary outcomes measures:** The primary outcome was mortality during the first 21 days following admission, secondary outcomes consisted of ICU-admission and ICU-mortality. Kaplan-Meier and Cox-regression analyses were used to determine the association with CVD risk factors.

**Results:** We included 1604 patients with a mean age of 66±15 of whom 60.5% were men. Antihypertensives, lipid lowering therapy, and antidiabetics were used by 45%, 34.7%, and 22.1% of patients. After adjustment for age and sex, the presence of ≥2 risk factors was associated with increased mortality risk (HR 1.52, 95%CI 1.15-2.02), but not with ICU-admission. Moreover, the use of ≥2 antidiabetics and ≥2 antihypertensives was associated with mortality independent of age and sex with HRs of respectively 2.09 (95%CI 1.55-2.80) and 1.46 (95%CI 1.11-1.91).

**Conclusions:** The accumulation of hypertension, dyslipidemia and diabetes leads to a stepwise increased risk for short-term mortality in hospitalized COVID-19 patients independent of age and sex. Further studies investigating how these risk factors disproportionately affect COVID-19 patients are warranted.

**Strengths and limitations of this study:**

- While previous data reported a high prevalence of CVD risk factors in COVID-19 patients, this study investigated whether diabetes, dyslipidemia and hypertension predict adverse outcomes.
- This study is limited by the use of medication as surrogate for cardiovascular risk factors
- The causality of the investigated risk factors remains to be addressed in future studies.

## Introduction

The global spread of coronavirus disease 2019 (COVID-19), first identified in Wuhan, China, in December 2019, has ignited an unprecedented ongoing global pandemic.(1) Although most infected individuals experience only mild symptoms that do not require hospitalization, the absolute number of patients requiring hospital admission is staggering. Risk stratification of these patients is crucial to optimize the use of hospital resources.(2) Several associations with adverse outcomes in COVID-19 patients have been identified, including factors that also predispose to cardiovascular disease (CVD), such as older age, male sex, hypertension, overweight and diabetes.(3,4) Furthermore, individuals with overt CVD appear to be affected more seriously by COVID-19 infection.(5)

The association between cardiovascular events and infectious diseases is well established. Examples include the increased prevalence of myocardial infarction during influenza pandemics,(6) and the higher number of cardiac complications in patients hospitalized for community-acquired pneumonia.(7) However, there are conflicting data whether the presence of shared CVD and COVID-19 risk factors merely reflect advanced age and history of ischemic heart disease in patients who develop severe infection, or are independently associated with adverse outcomes in the COVID-19 patient population.(4,8) For example, a higher than expected prevalence of diabetes, hypertension, obesity, and history of CVD was reported during the previous of outbreak Middle East respiratory syndrome coronavirus (MERS-CoV), which shares many similarities with COVID-19.(9)

In the present study, we hypothesized that three major risk independent CVD risk factors are associated with adverse outcomes in COVID-19 patients. To this end, we evaluated the time-to-event relationship between COVID-19 disease outcomes and a history of medication use for hypertension, dyslipidemia, and diabetes mellitus in a large prospective Dutch cohort of hospitalized COVID-19 patients.

## Patients and Methods

### Study design

COVID-PREDICT is a Dutch multicenter initiative to collect data of hospitalized patients with confirmed COVID-19. For this study, patients from 8 participating hospitals, including two university hospitals were included. All hospitalized patients >18 years with a positive COVID-19 polymerase chain reaction (PCR) or high suspicion based on CT-imaging of the thorax were included.(10) A waiver for the use hospital record data was obtained from the Medical Ethical Committees of the participating centers. Patients were given the opportunity to opt out.

The collected data was updated with daily reports on vital signs, laboratory results, complications, and clinical outcomes. In addition, the use of antihypertensive, lipid-lowering, and/or antidiabetic medication was determined from the home medication list. These were used as surrogates for hypertension, dyslipidemia, and diabetes. Antihypertensive medication was categorized as using either 0, 1, more than 1 of the following categories: non-dihydropyridine calcium channel blockers (CCBs), renin angiotensin system (RAS)-inhibitors (either angiotensin receptor antagonist or angiotensin receptor blockers) and diuretics (either loop diuretics, thiazide or thiazide like diuretics or potassium channel blockers). Lipid-lowering therapy was classified as the use of statin, ezetimibe, fibrates, or PCSK9-inhibitors. Antidiabetic medication was classified as using either 0, 1 or more than 1 of the following classes: metformin, sulfonylurea derivates, GLP-1 agonists, DDP4-inhibitors, SGLT-2 inhibitors, and insulin. Obesity was defined as a BMI >30 kg/m. Smoking was categorized into current or non/former smoker. The combined use of beta-blockers and platelet aggregation inhibitors was used as a surrogate for history of ischemic cardiac disease. Outcomes were determined 3 and 6 weeks after admission, or earlier when the patient died or was discharged from the hospital.

### Primary and secondary outcomes

The primary outcome consisted of overall mortality during the first 21 days following admission. Overall mortality was defined as either mortality during admission or discharge for palliative care, either at home or a palliative care facility. If the patient was discharged alive from the hospital and no further follow-up data was available, we considered the patient to be event free for the whole study period. Secondary outcomes consisted of ICU-admission and mortality in the subset of patients who had been admitted to the ICU.

### Statistical analysis

For the analysis, we included all consecutive patients who were primarily admitted to one of the participating centers between February 27^th^ and July 4^th^ 2020. Patients with unknown medication use prior to hospitalization were excluded. Patients were categorized based on the presence of either 0, 1 or more than 1 medication based cardiovascular risk factor. Baseline characteristics were depicted as mean ± standard deviation for normally distributed data, nonnormally as median [interquartile range], or as number (percentage) for categorical variables, and were compared using the appropriate tests (ANOVA, Kruskal-Wallis, chi-squared). The relationship between outcomes and cumulative cardiovascular risk factors was determined using a Kaplan-Meier analysis. We then determined hazard ratios using Cox-regression after correction for age and sex, and with additional correcting for obesity, smoking and history of ischemic cardiac disease. For ICU-mortality, a landmark analysis was performed starting from ICU-admission. Next, with Cox-regression models using the same covariates we determined the association between mortality and the use of antihypertensive, lipid-lowering, and antidiabetic drugs. All statistical analyses were conducted with R version 3.6.3 (R Foundation, Vienna, Austria) using the Survival version 3.1-11 and Tableone version 0.11.1 packages.

## Results

### Patient characteristics

Between February 27^th^ 2020 and July 4^th^ 2020, a total of 1614 patients with a confirmed COVID-19 infection were primarily admitted to one of the participating centers in the Netherlands. After exclusion of patients with an unknown medication list, we included 1604 patients in the present analysis. Their mean age was 66±15 years, 67.6% were Caucasian, and 60.5% were men. 6.5% of the admitted patients were current smokers. The majority of admitted patients (924, 57.6%) used some form of cardiovascular medication prior to admission. Antihypertensive medication was used by 721 (45.0%) patients, of whom 497 (68.9%) used RAS-inhibitors, 256 (35.5%) calcium-antagonists, and 374 (51.9%) diuretics. Lipid-lowering therapy was used by 557 (34.7%) patients, predominantly consisting of statins (540, 96.9%). In total, 354 (22.1%) patients used antidiabetic medication, of whom 282 (79.7%) used metformin and 137 patients (38.7%) insulin. 167 patients (10.4%) used the combination of a beta-blocker and a platelet aggregation inhibitor, reflecting a history of ischemic cardiac disease. In the subset of 566 patients for whom a more detailed medical history was available, we found a similar prevalence of cardiovascular disease, with 13.1% of the patients having a history of coronary artery disease, 3.7% of heart failure, 7.4% of stroke and 1.6% of peripheral arterial disease (see Supplement 3).

### Cardiovascular risk factors as markers for mortality and ICU-admission

In the entire cohort, 308 (19.2%) of the patients died or were discharged for palliative care. In total 273 (17.0%) of the patients were admitted to the ICU, of whom 78 died. 1100 (68.6%) patients were discharged alive from the hospital, 50 (3.1%) were transferred to another hospital. The remaining patients were still admitted to the hospital at time of the data collection (126; 7.9%) or follow-up data was not available yet (20; 1.25%). The Kaplan-Meier analysis showed a significant association between cardiovascular risk factors and overall mortality (p<0.0001; Figure 1A) and a trend towards increased ICU-mortality (p=0.055; Supplement 1). We found no association between cardiovascular risk factors and ICU-admission (p=0.85; Figure 1B). In Cox-regression analysis, a 5-year age increase was associated with a HR of 1.37 (CI 1.31-1.45) for mortality, while there was no significant association with sex (HR 1.02, CI 0.81-1.28). The presence of two or more cardiovascular risk factors was significantly associated with overall mortality (HR 1.52, 95%CI 1.15-2.02), but not with ICU-admission or ICU-mortality (Table 2). After additional correction for smoking, obesity, and the combined use of beta-blockers and platelet aggregation inhibitors the presence of two or more risk factors remained associated with mortality (HR 1.38, 95%CI 1.02-1.86, Supplement 2).

**Table 1.** Baseline characteristics.

|  | Overall | 0 RF | 1 RF | ≥2 RF | p |
| --- | --- | --- | --- | --- | --- |
| n | 1604 | 680 | 400 | 524 |  |
| Age (mean (SD)) | 65.67 (15.06) | 58.59 (15.71) | 69.93 (13.01) | 71.62 (11.45) | <0.001 |
| Women | 633 (39.5) | 298 ( 43.8) | 170 (42.5) | 165 (31.5) | <0.001 |
| Chronic cardiac disease | 467 (29.2) | 71 ( 10.5) | 117 (29.3) | 279 (53.4) | <0.001 |
| Hypertension | 734 (46.0) | 80 ( 11.8) | 242 (60.8) | 412 (78.8) | <0.001 |
| Chronic pulmonary disease | 288 (18.0) | 92 ( 13.6) | 82 (20.6) | 114 (21.9) | <0.001 |
| Asthma | 179 (11.2) | 77 ( 11.4) | 41 (10.3) | 61 (11.7) | 0.793 |
| Chronic kidney disease | 150 ( 9.4) | 17 ( 2.5) | 48 (12.0) | 85 (16.3) | <0.001 |
| Diabetes | 411 (25.7) | 19 ( 2.8) | 66 (16.5) | 326 (62.3) | <0.001 |
| Malignant neoplasm | 98 ( 6.2) | 36 ( 5.3) | 25 ( 6.3) | 37 ( 7.1) | 0.455 |
| Chronic hematologic disorder | 57 ( 3.6) | 32 ( 4.7) | 11 ( 2.8) | 14 ( 2.7) | 0.1 |
| Smoking | 78 ( 6.5) | 30 ( 6.0) | 16 ( 5.4) | 32 ( 7.8) | 0.383 |
| Obesity | 464 (30.8) | 164 ( 26.1) | 112 (29.4) | 188 (37.8) | <0.001 |
| Combined use of beta-blockers and antiplatelet drugs | 167 (10.4) | 7 ( 1.0) | 35 ( 8.8) | 125 (23.9) | <0.001 |
| Antihypertensive-Rx |  |  |  |  | <0.001 |
| 0 | 883 (55.0) | 680 (100.0) | 145 (36.2) | 58 (11.1) |  |
| 1 | 377 (23.5) | 0 ( 0.0) | 141 (35.2) | 236 (45.0) |  |
| 2 | 282 (17.6) | 0 ( 0.0) | 95 (23.8) | 187 (35.7) |  |
| ≥ 3 | 62 ( 3.9) | 0 ( 0.0) | 19 ( 4.8) | 43 ( 8.2) |  |
| Lipid-lowering-Rx |  |  |  |  | <0.001 |
| 0 | 1047 (65.3) | 0 (0.0) | 296 (74.0) | 89 (13.5) |  |
| ≥ 1 | 557 (34.7) | 0 ( 0.0) | 104 (26.0) | 453 (86.5) |  |
| Glucose-lowering-Rx |  |  |  |  | <0.001 |
| 0 | 1250 (77.9) | 680 (100.0) | 359 (89.8) | 211 (40.3) |  |
| 1 | 170 (10.6) | 0 ( 0.0) | 23 ( 5.8) | 147 (28.1) |  |
| ≥ 2 | 184 (11.5) | 0 ( 0.0) | 18 ( 4.5) | 166 (31.7) |  |

RF, risk factor; SD, standard deviation; IQR, interquartile range; Rx, medication. P-values indicate comparison between subgroups based on the presence of cumulative risk factors.

**Table 2.** Effect of cumulative cardiovascular risk factors on primary and secondary outcomes.

|  | Cumulative risk factors | HR | 95%CI |  | p |
| --- | --- | --- | --- | --- | --- |
| Mortality | 0 RF | 1 (ref) |  |  |  |
|  | 1 RF | 1.04 | 0.76 | 1.43 | 0.786 |
|  | ≥2 RF | 1.52 | 1.15 | 2.02 | 0.004 |
| ICU admission | 0 RF | 1 (ref) |  |  |  |
|  | 1 RF | 1.11 | 0.80 | 1.53 | 0.534 |
|  | ≥2 RF | 1.15 | 0.85 | 1.56 | 0.355 |
| ICU-mortality | 0 RF | 1 (ref) |  |  |  |
|  | 1 RF | 0.86 | 0.46 | 1.60 | 0.625 |
|  | ≥2 RF | 1.52 | 0.90 | 2.55 | 0.115 |

Cox-regression for the effect of cumulative cardiovascular risk factors on mortality, ICU-admission and ICU-mortality, corrected for sex and age. HR, hazard ratio; CI, confidence interval; ICU, intensive care unit.

**Table 3.**
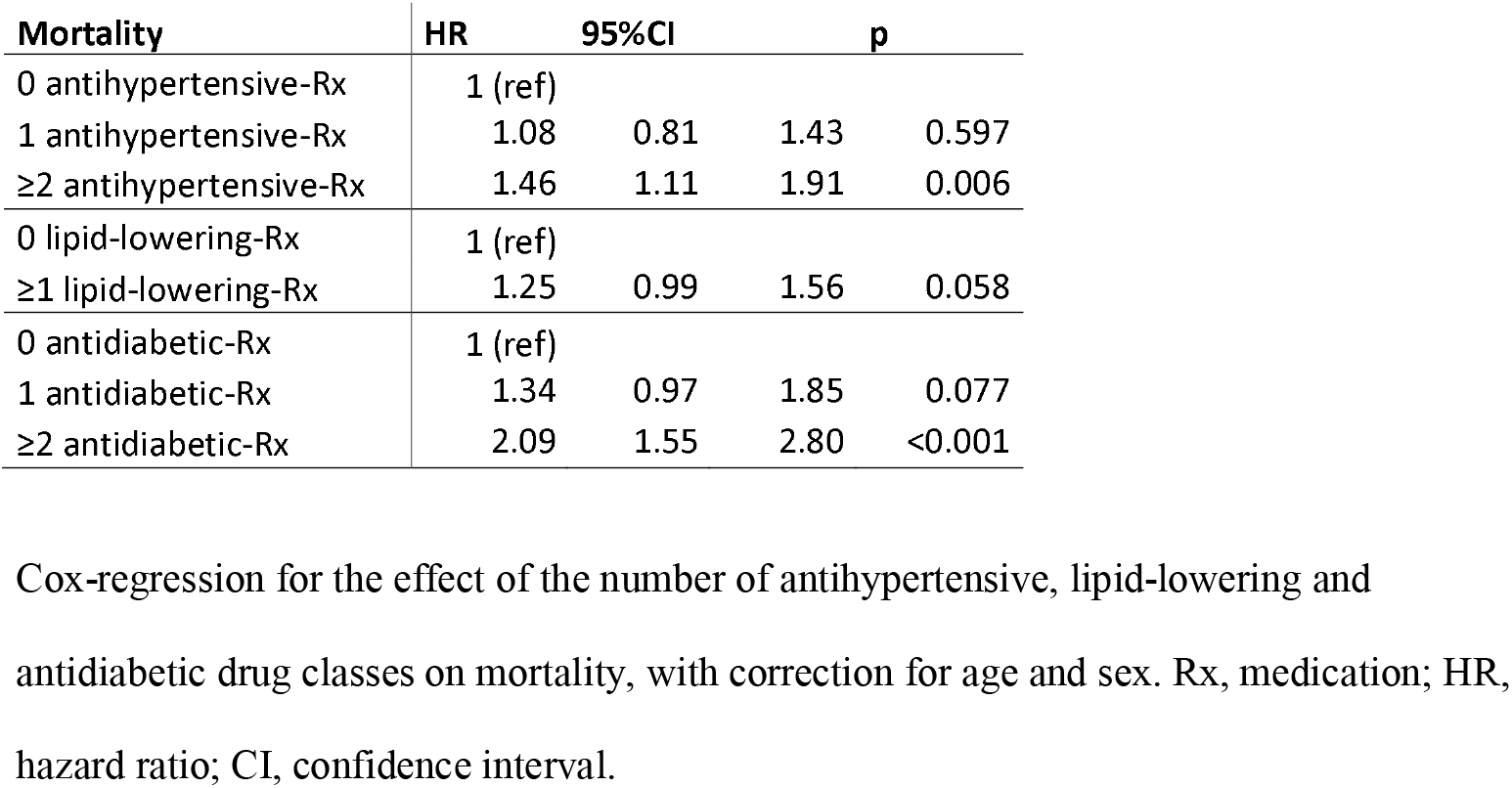
Effect of antihypertensive, lipid-lowering, and antidiabetic medications on mortality.

**Figure 1.**
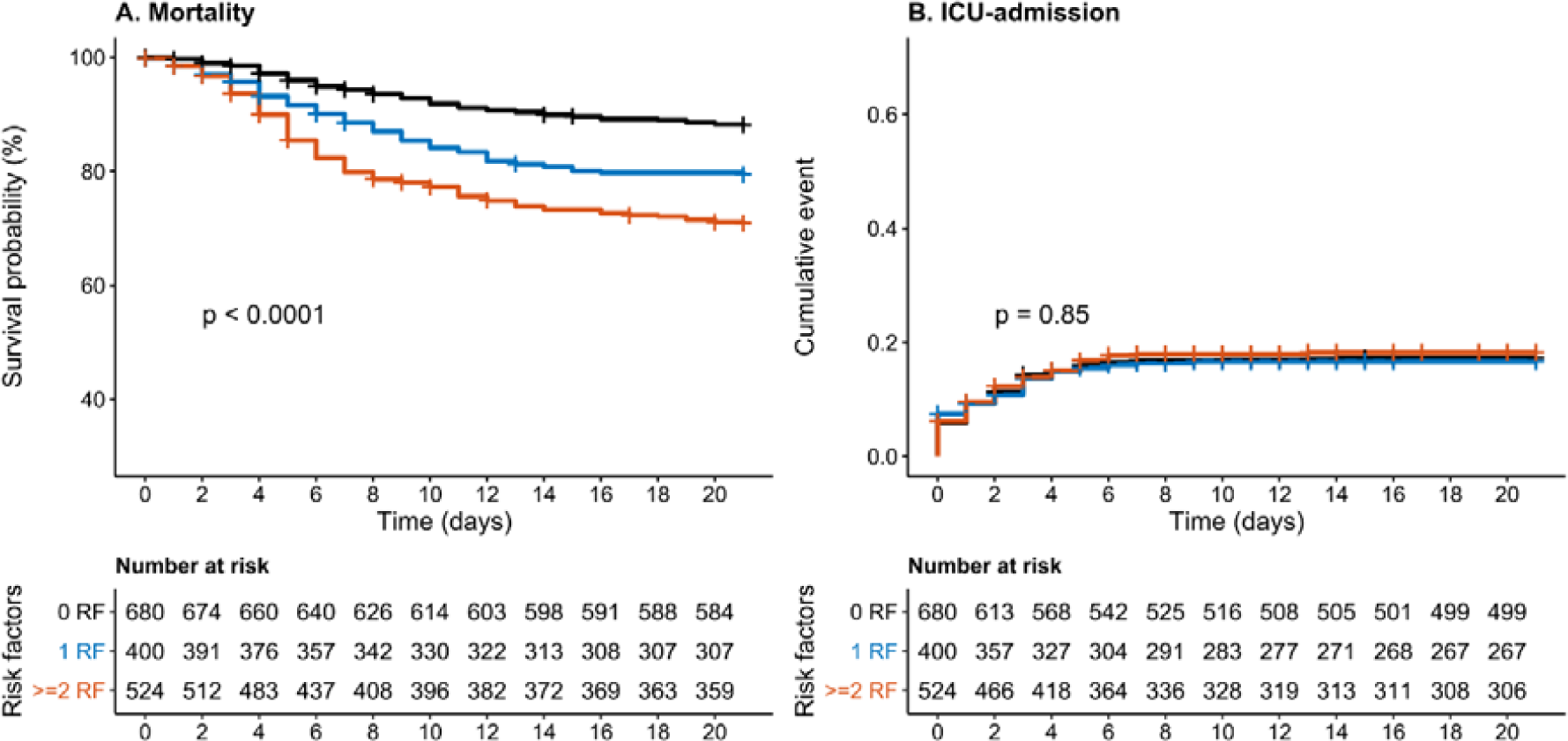
Survival and time-to event analysis of cumulative cardiovascular risk factors on mortality and ICU-admission. Kaplan-Meier analysis of hypertension, dyslipidemia and diabetes stratified into 0, 1 or more risk factors versus adverse clinical outcomes. Left panel (A) depicts mortality, right panel (B) ICU-admission. Log-rank test was used to test for differences between curves. RF, risk factor; ICU, intensive care unit.

### Individual risk factors

In the Cox-regression models corrected for age and sex, we observed that the use of two or more different classes of antihypertensives and antidiabetics were associated with 21-day mortality, with HR of respectively of 1.46 (95%CI 1.11-1.91) and 2.09 (95%CI 1.55-2.80). Similarly, we found a HR of 1.25 (95%CI 0.99-1.56) for the use of lipid-lowering medication. Additional correction for smoking, obesity and the combined use of beta-blockers and antiplatelet medication attenuated the association between the use of BP-lowering and lipid-lowering medication towards 1.33 (95%CI 1.01-1.76) and 1.14 (95%CI 0.89-1.45) for the use of ≥2 BP-lowering drugs or ≥1 lipid-lowering drug respectively. The association between the use of 2 or more glucose-lowering medications and mortality remained significant with an adjusted HR of 1.93 (95%CI 1.43-2.62; Supplementary table 2).

## Discussion

In a large Dutch cohort of hospitalized COVID-19 patients, we observed that patients with more than one risk factor for CVD had a 52% higher 3-week mortality risk, independent of age and sex. In addition, our data show that the use of two or more antihypertensives or antidiabetics, or one lipid-lowering drug is associated with adverse outcomes in COVID-19 patients. Patients using two or more antidiabetic drugs had the highest mortality risk. This suggests that patients with a history of or at high risk for cardiovascular disease have an increased risk for adverse COVID-19 disease outcomes.

The prevalence of medication use for CVD risk factors was higher in COVID-19 patients than in previously described cohorts representative for the general Dutch population of similar age.(11) Wuhan-based COVID-19 cohorts were the first to describe a higher mortality in those with hypertension and diabetes.(12) However, the average age and reported prevalence of CVD risk factors was much lower in these cohorts than in the present study.(4) The increased case-fatality rate in Europe and the US compared with China may in part be attributable to demographic differences in the COVID-19 infected population.(13) Recent European and US-based cohorts indeed demonstrate a higher average age with a similar distribution of hypertension and diabetes compared to our study, and also show a higher mortality in COVID-19 patients with hypertension and diabetes.(14,15) In line with the results from a large US-based cohort, we observed that age was significantly associated with mortality, while we found no significant difference in mortality between sexes.(16) This suggests that although men are more often hospitalized, there is no substantial difference in mortality after admission for severe COVID-19 infection. We add to these findings that the accumulation of CVD risk factors is associated with mortality, independent of age, sex, presence of coronary artery disease, smoking and obesity in hospitalized patients.

Analogous to COVID-19, CVD risk factors are also prevalent among patients hospitalized for community-acquired pneumonia.(17) However, we observed a stronger association between hypertension, diabetes, dyslipidemia and mortality in COVID-19 patients compared with previous studies on community-acquired pneumonia, despite a similar prevalence of CVD risk factors.(18) In addition, these effects remained significant after correction for covariates such as smoking, obesity, and the use of beta-blockers and antiplatelet drugs as surrogate for a history of ischemic cardiac disease. This might suggest that CVD risk factors in COVID-19 patients disproportionally affect the clinical course of COVID-19 patients compared to other infectious diseases. The present findings are comparable to the previous MERS-CoV outbreak, which also saw a preponderance of hypertension and diabetes in hospitalized patients.(9) Because both coronaviruses enter the cell through the angiotensin converting enzyme 2 (ACE2) receptor, a hypothesis has become that upregulation of ACE2 in patients with hypertension and/or diabetes facilitates transmission of the virus.(19–22) However, recent studies have shown no association between the use of RAS-medication and the disease course of COVID-19.(23,24) Furthermore, this theory does not explain the increased risk of mortality paired with other CVD risk factors in COVID-19 patients. An alternative explanation is that CVD risk factors predispose to myocardial injury in COVID-19 infected patients, contributing to a more severe clinical course.(25) Two recent studies revealed viral RNA in the myocardium of COVID-19 patients, suggesting that SARS-CoV-2 might infect the heart directly. It can be speculated that those with (pre-clinical) atherosclerosis are prone to experience coronary ischemia from viral myocardial involvement.(26,27)

In our cohort of COVID-19 patients, the presence of diabetes had the strongest association with mortality and remained significant after correction for covariates such as smoking, obesity, and the use of beta-blockers and antiplatelet drugs as surrogate for coronary artery disease. We observed smaller effect sizes for the presence of hypertension and dyslipidemia. These findings might suggest that diabetes predisposes for adverse outcomes in COVID-19 patients not only through its association with CVD, but potentially via other pathophysiological pathways specific to diabetes. A recent Chinese cohort showed diabetes to be associated with higher ICU-admission and more in-hospital mortality, but not independently of hypertension and history of CVD.(8) Interestingly, in those with diabetes, hypertension was associated with in-hospital death, independently of history of CVD, further supporting the additive effect of CVD risk factors on COVID-19 mortality. Nevertheless, it is hard to disentangle the precise relation based on epidemiological data. In line with current guidelines on CVD risk management, cardiovascular medication from different classes were often prescribed together in our cohort, also in patients with diabetes.(28) This makes it difficult to assess their separate contribution to mortality. It remains, therefore, unknown whether diabetes alone is associated with a higher risk of adverse outcomes or whether it is merely a reflection of increased vascular ageing in combination with the other risk factors.(29) In contrast to our results, the recent cohort of Cummings et al, found that among cardiovascular risk factors, only chronic cardiac disease was a strong predictor for hospital mortality, while smaller associations were found for other risk factors, including diabetes.(16) In line, a recent Italian cohort of hospitalized COVID-19 patients showed an increased prevalence of hypertension and diabetes amongst non-survivors, but only diabetes was an independent predictor after correction for other comorbidities.(30) In the present study, the cumulative presence of CVD risk factors did not show an association with increased risk for ICU-admission. This may have been influenced by selection prior to ICU admission, where the presence of co-morbidity was taken into account in the shared decision-making process, leading to a relative underrepresentation of patients with CV-risk factors.

The present analysis has several limitations. First, data collection was based on data collection forms of the WHO, which did not include detailed information on cardiovascular disease history. For this reason, we relied on medication use as a surrogate marker for established cardiovascular risk factors or disease, which has been used before in big cohort studies.(31) Nevertheless, some of these drugs might have been prescribed for different indications. Secondly, we only obtained follow-up during the first 21 days, however as depicted in the Kaplan-Meier analysis, almost all events occurred during the first 14 days, in line with earlier descriptions.(4) Finally, we cannot exclude that mortality in the current study is partially caused by other factors than COVID-19. However, as we used 21-day mortality as our primary outcome and only included patients admitted to the hospital with confirmed COVID-19 infection, it is very likely that the majority of deaths were directly attributable to COVID-19.

In conclusion, the accumulation of CVD risk factors leads to a stepwise increased risk for short-term mortality in hospitalized COVID-19 patients. Patients with diabetes had the highest risk, followed by similar risks for hypertension and dyslipidemia. Mechanistic studies investigating how CVD risk factors disproportionately affect COVID-19 patients compared to other infectious diseases are warranted.

## Data Availability

The data that support the findings of this study are available from the corresponding author, upon reasonable request.

## Acknowledgements

We would like to thank the CovidPredict consortium (www.covidpredict.org) for their efforts in providing the patient data.

## Author contributions

DC, NN, YK, LR, ES, BjvdB, LV, MB, PE conceptualized and designed the study. DC, NN, YK, LR performed the data analysis. DC, NN, YK, ES, BjvdB drafted the manuscript. All authors have made substantial contributions to the following: (1) the conception and design of the study, or acquisition of data, or analysis and interpretation of data, (2) revising the manuscript critically for important intellectual content, (3) final approval of the version to be submitted.

## Funding

DC is supported by a ZonMW grant (project number: 10430022010002).

## Competing interests

N.S.N. and L.F.R. are co-founders of Lipid Tools. E.S.G.S. reports personal fees from Amgen, personal fees from Sanofi-Regeneron, personal fees from Esperion, grants from Athera, outside the submitted work.

## Supplement 1. Survival analysis for ICU-mortality

**Figure S1.**
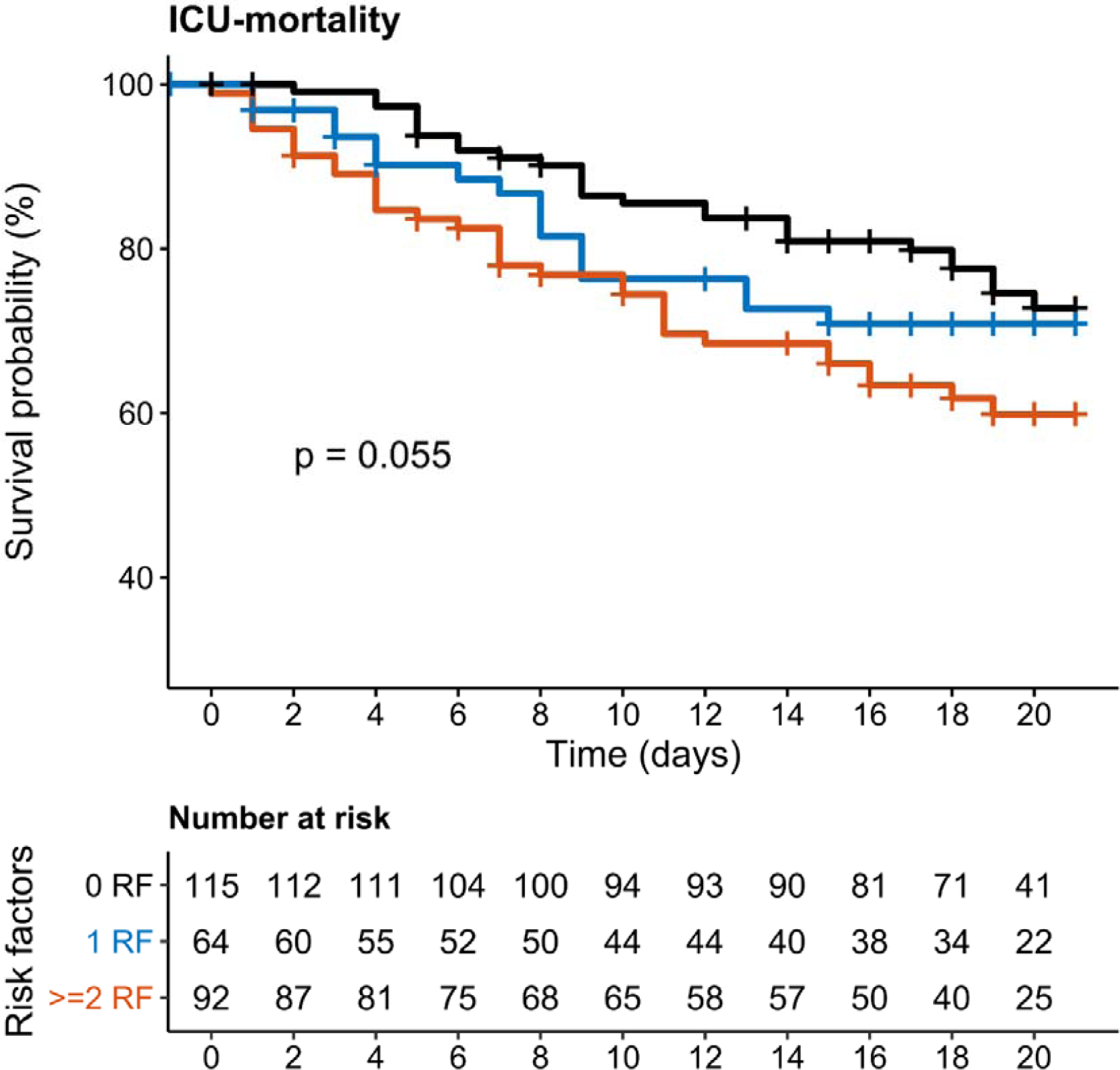
Survival analysis of cumulative cardiovascular risk factors for ICU-mortality. Kaplan-Meier analysis of hypertension, dyslipidemia and diabetes stratified into 0, 1 or more risk factors versus adverse clinical outcomes for mortality in patients admitted to the ICU. Log-rank test was used to test for differences between curves. RF, risk factor; ICU, intensive care unit.

## Supplement 2: Cox-regression models with additional correction for smoking, obesity and the use of both a beta-blocker and antiplatelet drug

We performed an additional analysis for the association between the cardiovascular risk factors and mortality with correction for smoking, obesity and the use of both a beta-blocker and antiplatelet drug. Data for smoking status was missing for 22.7% of the patients, data for obesity was missing for 9.2% of the patients. Imputation for these covariates was performed using the Multivariate Imputation by Chained (mice) package version 3.8.0. The pooled results averaged over 25 iterations are depicted in Supplementary Table 2. A complete cases analysis showed similar results (data not shown).

**Supplementary table S1:**
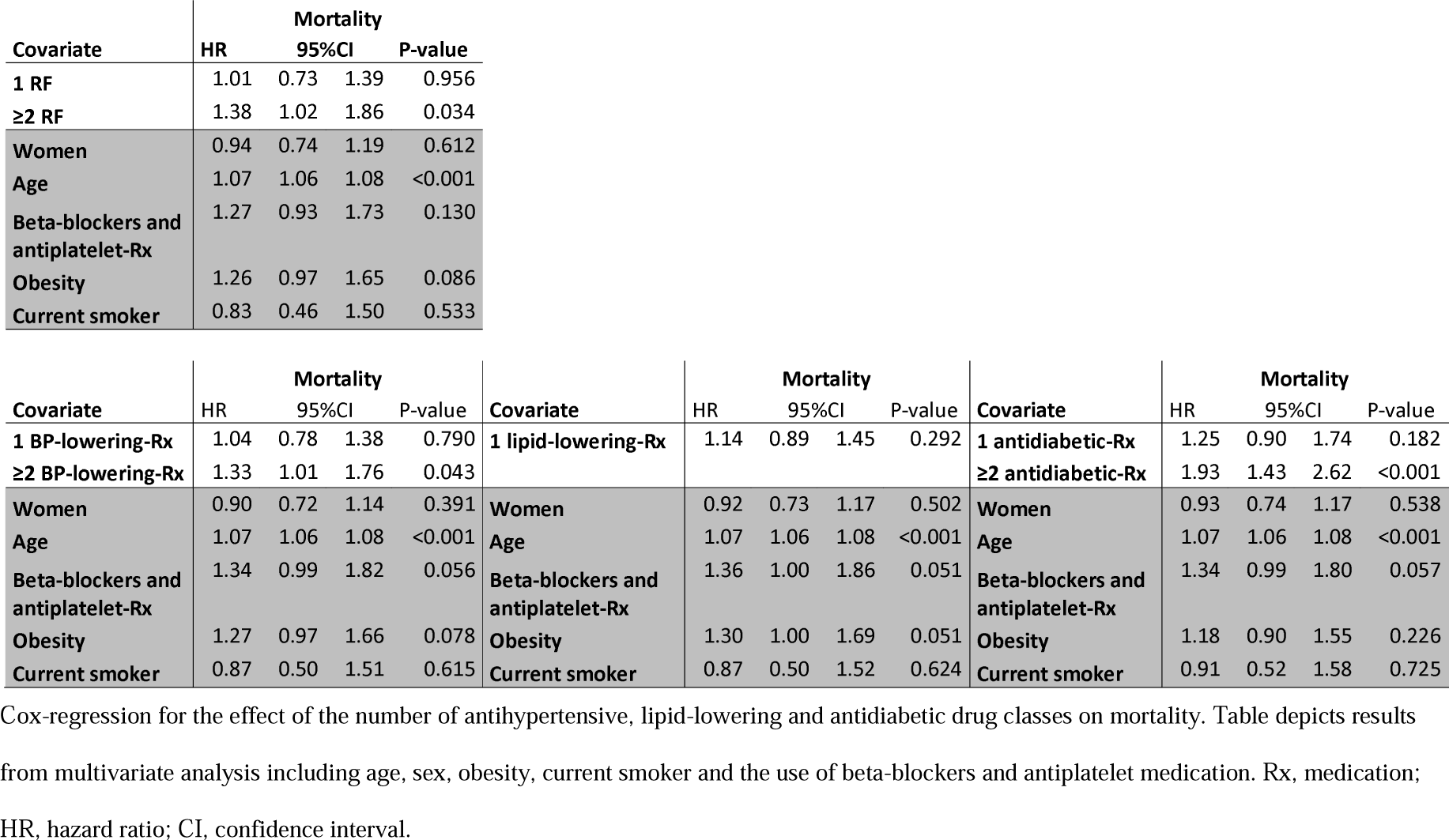
Effect of the cumulative risk factors, antihypertensive medication, lipid-lowering medication and antidiabetic medication on mortality after correction for covariates.

## Supplement 3: Baseline characteristics in subset with detailed clinical history

In three of the participating hospitals (Amsterdam UMC location AMC, Amsterdam UMC location VUMC, Flevoziekenhuis), we collected additional clinical information to validate the medication use as surrogate marker for cardiovascular risk factors and disease.

**Supplementary table S2:** Baseline characteristics of complete cohort, and subgroup where additional information about prior cardiovascular events was available.

| n | Overall<br>1604 | Subset<br>566 |
| --- | --- | --- |
| Age (mean (SD)) | 65.67 (15.06) | 61.37 (14.60) |
| Women | 633 (39.5) | 241 (42.6) |
| Chronic cardiac disease | 467 (29.2) | 125 (22.1) |
| Hypertension | 734 (46.0) | 252 (44.8) |
| Chronic pulmonary disease | 288 (18.0) | 77 (13.6) |
| Asthma | 179 (11.2) | 56 ( 9.9) |
| Chronic kidney disease | 150 ( 9.4) | 57 (10.1) |
| Diabetes | 411 (25.7) | 154 (27.2) |
| Malignant neoplasm | 98 ( 6.2) | 31 ( 5.5) |
| Chronic hematologic disorder | 57 ( 3.6) | 23 ( 4.1) |
| Smoking | 78 ( 6.5) | 28 ( 6.2) |
| Obesity | 464 (30.8) | 168 (32.8) |
| Combined use of beta-blockers and antiplatelet drugs | 167 (10.4) | 50 ( 8.8) |
| History of coronary artery disease |  | 74 (13.1) |
| History of heart failure |  | 21 ( 3.7) |
| History of stroke |  | 42 (7.4) |
| History of peripheral artery disease |  | 9 (1.6) |
| Antihypertensive-Rx |  |  |
| 0 | 883 (55.0) | 327 (57.8) |
| 1 | 377 (23.5) | 126 (22.3) |
| 2 | 282 (17.6) | 81 (14.3) |
| ≥ 3 | 62 ( 3.9) | 32 ( 5.7) |
| Lipid-lowering-Rx |  |  |
| 0 | 1047 (65.3) | 399 (70.5) |
| ≥ 1 | 557 (34.7) | 167 (29.5) |
| Glucose-lowering-Rx |  |  |
| 0 | 1250 (77.9) | 435 (76.9) |
| 1 | 170 (10.6) | 60 (10.6) |
| ≥ 2 | 184 (11.5) | 71 (12.5) |

RF, risk factor; SD, standard deviation; IQR, interquartile range, Rx medication.

## Notes

### Author Declarations

The institutional review board of the Amsterdam UMC, Amsterdam, the Netherlands

